# Computational modelling of decision-making in suicide attempters

**DOI:** 10.64898/2026.10.09.26365056

**Authors:** Camille Grandé, Lucas De Zorzi, Mocrane Abbar, Gerd Wagner, Fabrice Jollant

**Author notes:** Corresponding author: Pr. Fabrice Jollant, Hôpital Paul Brousse, Pôle de psychiatrie et addictologie, 12 avenue Paul Vaillant-Couturier 94800 Villejuif, France.

## Abstract

**Importance:** Suicide is a leading cause of mortality, yet its prevention remains a challenge. Suicide attempters exhibit disrupted decision-making, but the specific cognitive processes driving this impairment and their utility for risk stratification are unclear.

**Objective:** To determine which computational decision-making mechanisms are disrupted in high-risk suicide attempters, and whether they distinguish attempters from depressed and healthy controls.

**Design:** This cross-sectional study fit reinforcement-learning models to Iowa Gambling Task performance across two independent studies: SPAD (August 2017 to May 2020); SUICIDE-DECIDE (March 2021 to September 2023). Data was analyzed between January 2025 and July 2026.

**Setting:** SPAD was conducted in Nîmes (France). SUICIDE-DECIDE was a muticenter study across Jena (Germany) and Nîmes. Clinical groups were recruited from academic inpatient psychiatry units and outpatient clinics, and healthy controls through local advertisement.

**Participants:** SPAD included adults aged 60 years or older; SUICIDE-DECIDE middle-aged adults stratified by attempt means (violent/nonviolent). Consecutive participants were recruited into three groups: suicide attempters, patient controls, healthy controls.

**Main outcomes and measures:** The primary outcomes were group differences on parameters reflecting learning from rewards and losses, sensitivity to win frequency, choice perseverance, and forgetting of past choices. We trained a multinomial logistic regression model in the SUICIDE-DECIDE study using self-reported depression and parameters that differentiated attempters from controls across studies, and evaluated its performance in the SPAD sample.

**Results:** SPAD included 57 participants (40 females [70.2%]; mean [SD] age, 70.1 [8.2]). SUICIDE-DECIDE included 124 participants (77 females [62.1%]; 32.1 [11.0]). Suicide attempters forgot past choices faster than controls (SPAD: *F*_(2,36)_ = 13.5; *P* < .001; SUICIDE-DECIDE: *F*_(3,57)_ = 46.6; *P* < .001). No other parameter specifically differed in attempters across studies. In SUICIDE-DECIDE, greater forgetting of past choices was associated with attempter status only (RRR = 4.2, 95% CI [1.5, 11.5]). This parameter paired with depression severity correctly classified 87.2% of participants in SPAD (95% CI [74.3%, 95.2%], 41/47).

**Conclusions and relevance.:** Short-sighted decision-making may represent a cognitive phenotype associated with suicide in high-risk populations and a potential target for prevention and treatment.

**Key points:** *Question:* Which computational decision-making mechanisms are specifically disrupted in high-risk suicide attempters, and can they improve discrimination from depressed and healthy controls?

*Findings:* Across two independent case-control studies, suicide attempters consistently showed significantly greater forgetting of past choices. This computational parameter improved participant classification compared to depression severity alone, achieving a balanced accuracy in an independent validation sample of 100% for healthy controls, 85.95% for depressed controls, and 85.44% for suicide attempters.

*Meaning:* Greater forgetting of past choices may represent a reproducible cognitive phenotype associated with suicide risk that could improve risk stratification and targeted preventive interventions.

## Introduction

Suicide, one of the leading causes of mortality worldwide, claims more than 720,000 lives each year.^1^ Depression is one of the strongest risk factors for suicide,^2^ yet has limited predictive value, as nearly 90% of individuals with depression do not die by suicide.^3^ Identifying additional mechanisms that shape the transition from suicidal ideation to action remains a challenge.

Growing evidence points to cognitive impairments in suicide attempters (SA), especially riskier decision patterns on the Iowa Gambling Task (IGT).^4,5^ This task assesses how individuals balance immediate rewards against long-term losses through trial-and-error learning.^6,7^ Individuals who use a violent method (vSA) to end their life perform even worse.^8,9^ vSA are closer to suicide decedents in their clinical, sociodemographic and biological profiles^10,11^ and face higher suicide-death risk than SA who only used non-violent methods (nvSA).^12^ First-degree relatives of suicide decedents with no history of attempts also make riskier choices,^13,14^ pointing to a heritable trait rather than a suicide-related state.

Decision-making deficits can arise from multiple underlying mechanisms (*e.g.,* sensitivity to rewards/losses, past choices memory) which global IGT scores cannot isolate, potentially explaining inconsistent findings.^15^ Computational modeling uses mathematical equations that map actions (*e.g*., choice at time *t*) to future behavior (choice at time *t*_+1_) to infer the values of latent variables—the internal processes we cannot observe but assume operate in the brain—and compare them across groups.^16^

Computational parameters of the IGT show stronger reliability and sensitivity to individual differences than global scores,^17^ but remain underused in suicide research.

Using the Prospect Valence Learning–Delta model (PVL–Delta),^18^ Alacreu-Crespo et al^19^ found lower loss aversion and higher forgetfulness of past trials in SA compared to healthy controls (HC), but no differences emerged relative to depressed patient controls (PC).

The Outcome Representation Learning model (ORL)^20^ is a competing model which separates four strategies consistently observed in the IGT: maximizing long-term gains, maximizing wins, persevering on previous choices, and updating behavior depending on outcomes (*i.e.*, reversal learning). Using this model, Moret et al.^21^ reported that greater forgetfulness of past choices and stronger deck perseverance were associated with suicide attempt among individuals reporting high childhood trauma. They could not identify clear differences between SA and PC, likely because they treated SA as a homogeneous group despite known heterogeneity in intent, methods, and lethality, among others.^22^

To address this limitation, we studied two high-lethality risk phenotypes: older adults, who have the highest lethality rates per attempt in Western countries,^23^ and vSA, as explained above. We applied validated models of the IGT to identify latent mechanisms associated with suicidal behavior that operate independently of demographic or other clinical variables, thereby offering potential targets for cognition-based prevention and treatment. We further examined whether these parameters combined with depression severity predicted group membership, providing preliminary evidence for their utility in longitudinal prediction of suicidal behavior.

We hypothesized that older SA and vSA would exhibit differences on computational parameters of the IGT distinct from HC and PC, and that robust differences would increase participant classification accuracy compared to depression severity alone. We defined robust differences as replication of the same mechanism and direction of effect across the two samples. Findings observed in one sample are reported as exploratory and interpreted as less conclusive.

## Methods

This study leverages data from two independent case-control, cross-sectional investigations of cognition in SA: SPAD (older adults) and SUICIDE-DECIDE (stratification by attempt-method violence). Local ethics committees approved the protocols in accordance with the Declaration of Helsinki. All participants provided written informed consent. Reporting follows the Strengthening the Reporting of Observational Studies in Epidemiology (STROBE) guidelines.^24^

### Participants and procedure

#### SPAD

We recruited 80 participants aged 60 years or more between 2017 and 2020 at Nîmes Academic Hospital, France: (1) Healthy controls with no history of suicide attempt or mental disorder (HC, *n* = 28); (2) Depressed patient controls with no history of suicide attempt (PC, *n* = 22); (3) Depressed patients with a history of suicide attempt (SA, *n* = 30).

SA and PC were hospitalized or recruited in outpatient clinics, and currently suffered from a moderate/severe major depressive episode (MDE) according to DSM-5 criteria, that could be part of a bipolar disorder (BD) or major depressive disorder (MDD). Suicide attempts were defined as « *any action performed with some intention of causing one’s own death* »,^25^ excluding aborted or interrupted acts and non-suicidal self-harm. HC were recruited through local advertisement. Further information on exclusion criteria for both studies is available in eMethods in Supplement.

Participants completed the Mini Mental State Examination^26,27^ and French National Adult Reading Test (fNART)^28^ to ensure similar intellectual abilities, the Beck Depression Inventory–13 items,^29^ and a randomized neuropsychological battery including the IGT.

#### SUICIDE-DECIDE

We recruited 132 adults aged 18-60 years across two sites (93 at Jena Academic Hospital, Germany; 39 at Nimes Academic Hospital, France), using the same *a priori* groups (HC: *n* = 49; PC: *n* = 34; SA: *n* = 49). Suicide attempts were defined as “*a self-initiated sequence of behaviors by an individual who, at the time of initiation, expected that the set of actions would lead to his or her own death*”,^30^ and further classified as violent (vSA, *n* = 22) or non-violent (nvSA, *n* = 27) using the ICD-10 codes X66-X82 (eMethods in Supplements). Controls had no personal or family history of suicidal behavior in first or second-degree relatives. Both patient groups experienced a current MDE as part of a MDD or BD.

Participants completed, among other tests, the Multiple Choice Vocabulary Intelligence Test^31^ (German) or the fNART^28^ (French), Beck’s Depression Inventory-II^32^ and the IGT. Participants were told that their payment would depend on task performance to promote realistic gambling and motivation, but all ultimately received €100 after the assessment.

### Iowa Gambling Task

In this computerized task, participants iteratively select cards from four decks and experience their associated monetary gains and losses.^6^ They are instructed to maximize their final amount knowing that some decks are better than others. Decks A and B yield higher wins but larger long-term losses (disadvantageous). Decks C and D yield smaller wins but smaller losses (advantageous). Decks B and D have a higher win frequency (90:10) than decks A and C (50:50). Participants must learn to favor the advantageous decks over the riskier ones through 100 card selections. Classic behavioral analyses appear in eSupplements.

### Computational modelling

We fitted three validated models using *R* (v4.4) and *hBayesDM* (v2.0.0.9)^33^: the Prospect Valence Learning model with decay reinforcement-learning rule (PVL-Decay),^18,34^ PVL-Delta,^18,34^ and ORL.^20^ The ORL model performed best in both samples (eMethods in Supplements). It includes five parameters. Reward (*A_rew_* ∈ [0, 1]) and Punishment learning (*A_pun_* ∈ [0, 1]) track how quickly participants update their behavior after wins or losses (higher values correspond to faster learning). Decay-rate (*K* ∈ [0, 5]) reflects memory for past outcomes (lower or higher values suggest individuals remember long or short histories respectively). Win-frequency perseverance (*B_F_* ∈ (−∞, +∞)) captures preference for decks with lower (*B_F_* < 0) or higher (*B_F_* > 0) win-frequency. Deck perseverance (*B_P_* ∈ (−∞, +∞)) reflects tendency to switch (*B_P_* < 0) or stay (*B_P_* > 0) with recent choices.

### Statistical analyses

We estimated group differences in parameter hyperdistributions using Welch’s ANOVA and Games-Howell post-hoc test (*rstatix* v1.1.0),^35^ adjusting p-values with Holm’s method. Decay-rate was the sole parameter that differed in SA only across both studies, therefore we tested if it could be explained by demographic, clinical, or cognitive indicators using backward stepwise selection (Akaike Information Criterion (AIC)-based elimination, *p* > .10) on an ordinary least squares model followed by multiple linear regression. We used Multiple Imputation by Chained Equations to impute missing data (*mice* v3.19, eMethods and eResults; *n* = 149 post-imputation).^36^ We entered predictors (Group, age, sex, BDI z-score, Go/No-Go reaction time (RT), correct responses, omissions, commissions) and applied backward stepwise selection, retaining predictors with the strongest contribution. The final formula included: *K ∼ Sex + Group + Go/No-Go mean RT + Sample*, using robust HC3 corrections to account for heteroscedasticity (eResults in Supplements).

We used Multinomial Logistic Regression (*nnet* v7.3.20)^37^ to assess if depression severity (BDI total) and *K*, both z-scored, could predict group membership.

SUICIDE-DECIDE was used as the training sample due to larger size (*n* = 107 with complete BDI data). We compared two models (1: Group ∼ BDI; 2: Group ∼ BDI + *K*) using a likelihood ratio test and AIC to evaluate whether adding *K* improved prediction of group membership. We found no evidence of multicollinearity between the two predictors (*VIF* = 1.1). We evaluated model performance using classification accuracy, Cohen’s *κ*, per-class sensitivity, specificity, positive and negative predictive values (*caret* v7.0.1),^38^ and individual coefficient significance using Wald z-tests. We report exponentiated coefficients and their 95% confidence intervals as Relative Risk Ratios (RRR),^39^ using HC as the reference. Model 2 was applied to SPAD (*n* = 47) as a validation sample, and we compared performance to assess generalization.

## Results

### Population characteristics

SPAD included 57 participants with complete IGT data (Table 1): 22 HC, 15 PC, and 20 SA. SUICIDE-DECIDE included 124 participants with complete IGT data (Table 2): 44 HC, 34 PC, and 46 SA, among which 25 were considered nvSA, and 21 vSA.

**Table 1.** Demographics, Clinical and Cognitive Characteristics SPAD.

| Characteristic | Healthy controls | Patient controls | Suicide attempters | Missing | *P*-value <sup>a</sup> | Post-hoc <sup>b</sup> |
| --- | --- | --- | --- | --- | --- | --- |
| *n* with IGT data | 22 | 15 | 20 | N/A | N/A | N/A |
| Sociodemographic |  |  |  |  |  |  |
| Age (mean [SD]) | 66.4 [4.7] | 71.7 [8.1] | 73.1 [9.8] | 0 | .04 | HC < SA |
| Sex (No. Female [%]) | 12/22 [55%] | 13/15 [87%] | 15/20 [75%] | 0 | .09 | N/A |
| Education (No. [%]) |  |  |  |  |  |  |
| < *Secondary* | 3/19 [16%] | 1/13 [8%] | 3/19 [16%] | 6<br>(HC: 3,<br>PC: 2, SA: 1) | .41 | N/A |
| *Secondary school* | 6/19 [32%] | 7/13 [54%] | 11/19 [58%] |  |  |  |
| *High school* | 5/19 [26%] | 5/13 [38%] | 4/19 [21%] |  |  |  |
| *Bachelor* | 2/19 [11%] | 0/13 [0%] | 0/19 [0%] |  |  |  |
| *Master* | 2/19 [11%] | 0/13 [0%] | 0/19 [0%] |  |  |  |
| *Doctorate* | 1/19 [5%] | 0/13 [0%] | 1/19 [5%] |  |  |  |
| Clinical |  |  |  |  |  |  |
| MMSE (mean [SD]) | 28.8 [1.6] | 26.5 [1.5] | 27.3 [1.4] | 1 (SA) | < .001 | HC > PC, SA |
| fNART (mean [SD]) | 33 [4.3] | 30 [7.7] | 28.5 [8.4] | 0 | .13 | N/A |
| BDI-13 (mean [SD]) | 0.8 [1.0] | 10.5 [5.6] | 11.7 [6.1] | 10 (HC: 5, PC: 3, SA: 2) | < .001 | HC > PC, SA |
| SIS Total (mean [SD]) | N/A | N/A | 11.4 [7.1] | 3 | N/A | N/A |
| Suicide attempts past year (No. [%]) |  |  |  |  |  |  |
| 1 | N/A | N/A | 13/16 [81%] | 4 | N/A | N/A |
| 2 |  |  | 2/16 [13%] |  |  |  |
| 3 |  |  | 1/16 [6%] |  |  |  |
| Lethality most lethal attempt (No. [%]) |  |  |  |  |  |  |
| 0 | N/A | N/A | 1/16 [6%] | 4 | N/A | N/A |
| 1 |  |  | 4/16 [25%] |  |  |  |
| 2 |  |  | 5/16 [31%] |  |  |  |
| 3 |  |  | 2/16 [12%] |  |  |  |
| 4 |  |  | 4/16 [25%] |  |  |  |
| Cognition |  |  |  |  |  |  |
| IGT Total Score (mean [SD]) | −8.4 [20.1] | 12.3 [24.6] | −4.6 [22.3] | 0 | .51 | N/A |
| Go / No-Go (mean [SD]) |  |  |  |  |  |  |
| RT (ms) | 417 [68.5] | 481.2 [62.9] | 474.4 [69.9] | 1 (HC) | .003 | HC < PC, SA |
| Correct | 19.5 [1.4] | 19.6 [0.7] | 18.9 [2.1] |  | .40 | N/A |
| Commissions | 1.6 [1.6] | 1.7 [1.3] | 4.4 [3.2] |  | .005 | HC, PC < SA |
| Omissions | 0.5 [1.4] | 0.4 [0.7] | 1.2 [2.1] |  | .40 | N/A |
| Verbal Fluency (mean [SD]) |  |  |  |  |  |  |
| Phonemic | 17.8 [4.8] | 14.9 [4.1] | 12.6 [5.6] | 6 (HC: 2, PC: 1, SA: 3) | .02 | HC > SA |
| Categorical | 17.9 [7.9] | 12 [5.4] | 11.2 [6.3] |  | .007 | HC > PC, SA |
| Medication |  |  |  |  |  |  |
| (No. Yes [%]) |  |  |  |  |  |  |
| Anti Anxiolytic | 0/22 [0%] | 0/12 [0%] | 1/19 [5%] | 4 (PC: 3, SA: 1) | 40 | N/A |
| Antidepressant | 0/22 [0%] | 11/12 [92%] | 18/19 [95%] |  | < .001 | HC < SA, PC |
| Antipsychotic | 0/22 [0%] | 7/12 [58%] | 13/19 [68%] |  | < .001 | HC < SA, PC |
| Benzodiazepines | 0/22 [0%] | 8/12 [67%] | 15/19 [79%] |  | < .001 | HC < SA, PC |

Abbreviations: MMSE, Mini-Mental State Examination; fNART, french New Adult Reading Test; BDI-13, Beck Depression Inventory–13 items; SIS, Suicide Intent Scale; IGT, Iowa Gambling Task; RT, Reaction time.
<sup>a</sup> Groups were compared using the Chi-Square test for categorical variables, and Kruskal-Wallis test for continuous variables (eTable 2 in Supplement)
<sup>b</sup> Post-hoc comparisons were computed using proportion tests for categorical variables, and Dunn's test for numeric variables (eTable 2 in Supplement)

**Table 2.** Demographics, Clinical and Cognitive Characteristics SUICIDE-DECIDE.

| Characteristic | Healthy controls | Patient controls | Non violent suicide attempts | Violent Suicide attempts | Missing | P-value<sup>a</sup> | Post-hoc<sup>b</sup> |
| --- | --- | --- | --- | --- | --- | --- | --- |
| *n* with IGT data | 44 | 34 | 25 | 21 | N/A | N/A | N/A |
| **Sociodemographic** |  |  |  |  |  |  |  |
| Age (mean [SD]) | 31.9 [9.3] | 33.6 [11.6] | 31.8 [12.8] | 30.3 [11.8] | 0 | .54 | N/A |
| Sex (No. Female [%]) | 22/44 [50%] | 23/34 [68%] | 20/25 [80%] | 12/21 [57%] | 0 | .08 | N/A |
| Education (No. [%]) |  |  |  |  |  |  |  |
| *Special education school* | 0/35 [0%] | 0/29 [0%] | 0/23 [0%] | 2/21 [10%] | 16 (HC: 9, PC: 5, nvSA: 2) | .004 | HC, PC > nvSA |
| *Lower Secondary school* | 1/35 [3%] | 1/29 [3%] | 2/23 [9%] | 2/21 [10%] |  |  |  |
| *Intermediate Secondary school* | 3/35 [9%] | 5/29 [17%] | 12/23 [52%] | 4/21 [19%] |  |  |  |
| *Academic Secondary school* | 9/35 [26%] | 9/29 [31%] | 4/23 [17%] | 6/21 [29%] |  |  |  |
| *University* | 19/35 [54%] | 10/29 [34%] | 5/23 [22%] | 7/21 [33%] |  |  |  |
| *University of applied sciences* | 3/35 [9%] | 4/29 [14%] | 0/23 [0%] | 0/21 [0%] |  |  |  |
| **Clinical** |  |  |  |  |  |  |  |
| fNART (mean [SD])<sup>c</sup> | 25.8 [3.9] | 24 [3.5] | 25.5 [4.1] | 22.5 [3.5] | 0 | .50 | N/A |
| MWT-IQ (mean [SD])<sup>c</sup> | 103.7 [9.3] | 107.6 [12.9] | 103.6 [11.1] | 112.2 [17.2] | 3 (HC: 1; PC: 3) | .39 | N/A |
| BDI-II (mean [SD]) | 2.4 [4.0] | 24.2 [12.6] | 31.9 [14.4] | 30.1 [14.5] | 18 (HC: 9, PC: 6, nvSA: 3) | < .001 | HC < PC, nvSA, vSA |
| SIS Total (mean [SD]) | N/A | N/A | 18.1 [4.6] | 17.8 [5.4] | 4 (nvSA: 3, vSA: 1) | .48 | N/A |
| Suicide attempts past year (No. [%]) |  |  |  |  |  |  |  |
| 0 | N/A | N/A | 1/22 [5%] | 1/21 [5%] | 3 (nvSA) | .20 | N/A |
| 1 |  |  | 15/22 [68%] | 10/21 [48%] |  |  |  |
| 2 |  |  | 4/22 [18%] | 4/21 [19%] |  |  |  |
| 3 |  |  | 1/22 [5%] | 1/21 [5%] |  |  |  |
| ≥4 |  |  | 1/22 [5%] | 5/21 [24%] |  |  |  |
| Lethality most lethal attempt (No. [%]) |  |  |  |  |  |  |  |
| 0 | N/A | N/A | 2/12 [17%] | 4/10 [40%] | 24 (nvSA: 13, vSA: 11) | .37 | N/A |
| 1 |  |  | 0/12 [0%] | 1/10 [10%] |  |  |  |
| 2 |  |  | 3/12 [25%] | 1/10 [10%] |  |  |  |
| 3 |  |  | 6/12 [50%] | 2/10 [20%] |  |  |  |
| 4 |  |  | 1/12 [8%] | 2/10 [20%] |  |  |  |
| Cognition |  |  |  |  |  |  |  |
| IGT Total Score (mean [SD]) | 14.7 [31.0] | 1.1 [30.6] | 5.4 [33.0] | 1.6 [33.3] | 0 | > .99 | N/A |
| Go / No-Go (mean [SD]) |  |  |  |  |  |  |  |
| RT (ms) | 476.0 [52.2] | 489.1 [59.9] | 463.1 [69.9] | 473.0 [74.2] | 22 (HC: 11, PC: 8, nvSA: 3) | .42 | N/A |
| Correct | 300.0 [10.2] | 298.0 [10.5] | 293.2 [12.9] | 292.0 [15.3] |  | .06 | N/A |
| Commissions | 15.9 [9.0] | 17.8 [10.1] | 23.1 [14.2] | 21.2 [11.7] |  | .15 | N/A |
| Omissions | 4.1 [6.8] | 4.2 [2.5] | 3.7 [4.3] | 6.9 [14.5] |  | .31 | N/A |
| Verbal Fluency (mean [SD]) |  |  |  |  |  |  |  |
| Phonemic | 24.1 [8.1] | 20.4 [5.2] | 22.7 [8.8] | 23 [8.2] | 19 (HC: 10, PC: | .42 | N/A |
| *Categorical* | 41.2 [7.8] | 37.6 [9.5] | 37.3 [9.4] | 39.7 [15.0] | 7, nvSA: 2) | .32 | N/A |
| **Medication** |  |  |  |  |  |  |  |
| (No. Yes [%]) |  |  |  |  |  |  |  |
| *Dopamine agonist* | 0/44 [0%] | 0/28 [0%] | 1/23 [4%] | 0/21 [0%] | 8 (PC: 6, nvSA: 2) | .26 | N/A |
| *Benzodiazepine* | 0/44 [0%] | 2/28 [7%] | 5/23 [22%] | 3/21 [14%] |  | .02 | HC < nvSA, vSA |
| *Lithium* | 0/44 [0%] | 1/28 [4%] | 1/23 [4%] | 1/21 [5%] |  | .58 | N/A |
| *NMDA antagonist* | 0/44 [0%] | 0/28 [0%] | 1/23 [4%] | 0/21 [0%] |  | .25 | N/A |

Abbreviations: fNART, french New Adult Reading Test; MWT-IQ, Multiple Choice Vocabulary Intelligence Test (Mehrfachwahl-Wortschatz-Test); BDI-II, Beck Depression Inventory–II; SIS, Suicide Intent Scale; IGT, Iowa Gambling Task; NMDA, N-methyl-D-aspartate; RT, Reaction time
<sup>a</sup> Groups were compared using the Chi-Square test for categorical variables, and Kruskal-Wallis test for continuous variables (eTable 3 in Supplement)
<sup>b</sup> Post-hoc comparisons were computed using proportion tests for categorical variables, and Dunn's test for numeric variables (eTable 3 in Supplement)
<sup>c</sup> French participants completed the fNART, German participants the MWT-IQ. Values displayed here are for the corresponding participants

### Computational modelling

In SPAD, we observed significant group differences on punishment (*A_pun_* ; *F*_2,23_ = 70.8, *P* < .001) and reward (*A_rew_*; *F*_2,23_ = 86.1, *P* < .001) learning, win-frequency perseverance (*B_F_* ; *F*_2,30_ = 19.7, *P* < .001), and decay-rate (*K*; *F*_2,36_ = 13.5, *P* < .001).

Post-hoc contrasts showed decreased reward learning in SA compared with HC (mean difference [MD] = –0.31, 95% CI [–0.39, –0.22]) and PC (MD = –0.12, 95% CI [–0.15, –0.09]), and in PC compared to HC (MD = –0.19, 95% CI [–0.28, –0.10]). SA also showed decreased punishment learning (HC-SA: MD = –0.07, 95% CI [–0.09, –0.05]; PC-SA: MD = –0.06, 95% CI [–0.09, –0.04]), reduced win-frequency perseverance (HC-SA: MD = –1.15, 95% CI [–1.91, –0.38]; PC-SA: MD = –1.24, 95% CI [–1.73, –0.75]), and increased decay-rate (HC-SA: MD = 0.35, 95% CI [0.07, 0.62]; PC-SA: MD = 0.55, 95% CI [0.29, 0.81]). Control groups did not differ on those parameters (*A_rew_*: MD = –0.01, 95% CI [–0.04, 0.02]; *B_F_* : MD = 0.10, 95% CI [–0.55, 0.74]; *K*: MD = –0.20, 95% CI [–0.44, 0.03]).

In SUICIDE-DECIDE, we observed significant group differences on punishment (*A_pun_* ; *F*_3,64_ = 6.9, *P* < .001) and reward (*A_rew_*; *F*_3,55_ = 37.8, *P* < .001) learning, deck perseverance (*B_P_* ; *F*_3,52_ = 3.0, *P* = .04), and decay-rate (*K*; *F*_3,57_ = 46.6, *P* < .001).

Post-hoc contrasts showed decreased punishment learning in vSA compared with HC (MD = –0.06, 95% CI [–0.10, –0.01]) and nvSA (MD = –0.04, 95% CI [–0.06, –0.01]), and in PC compared with HC (MD = –0.05, 95% CI [–0.09, 0]). No difference emerged between vSA and PC (MD = –0.01, 95% CI [–0.04, 0.02]). vSA exhibited reduced reward learning compared to all other groups (HC-vSA: MD = –0.15, 95% CI [–0.21, –0.09]; PC-vSA: MD = –0.10, 95% CI [–0.14, –0.05]; nvSA-vSA: –0.18, 95% CI [–0.25, –0.10]), and increased deck perseverance compared to nvSA (MD = 5.04, 95% CI [0.26, 9.81]). Finally, we found increased decay-rates in vSA compared with HC (MD = 0.97, 95% CI [0.71, 1.24]), PC (MD = 1.13, 95% CI [0.87, 1.39]), and nvSA (MD = 0.78, 95% CI [0.50, 1.07]), and nvSA compared with PC (MD = 0.35, 95% CI [0.12, 0.57]), with no differences between controls (MD = –0.15, 95% CI [–0.35, 0.04]).

Increased decay-rate was the only difference we observed solely in suicide attempters across the two studies (Figure 1). Therefore, we focused on this particular parameter for subsequent analyses.

**Figure 1.**
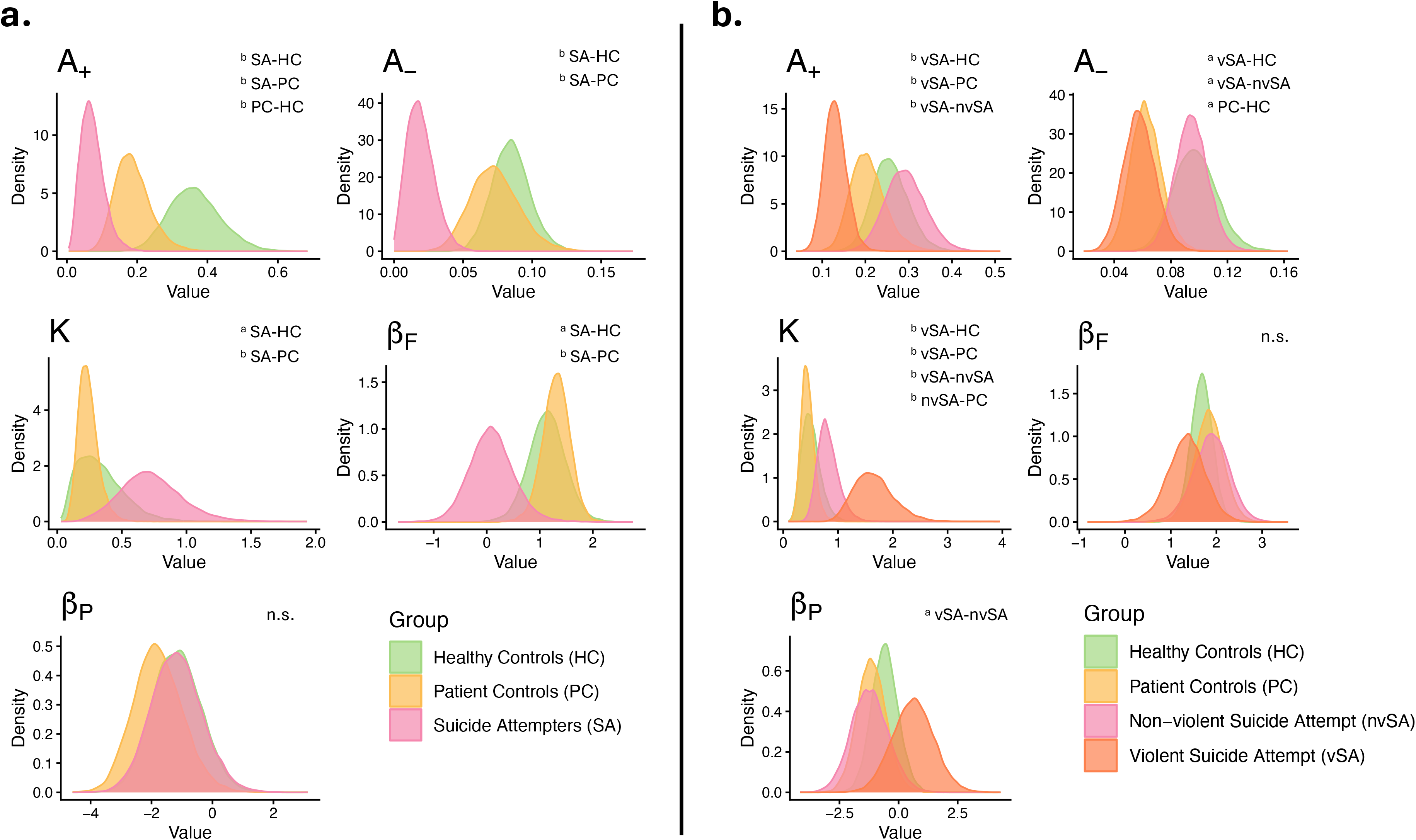
Outcome Representation Learning model parameters posterior estimates. Abbreviations: *A_rew_*, Reward learning rate (*A_rew_* ∈ [0, 1]); *A_pun_*, Punishment learning rate (*A_pun_* ∈ [0, 1]); *K*, Decay-rate (*K* ∈ [0, 5]); *B_F_*, Win-frequency perseverance (*B_F_* ∈ (−∞, +∞)); *B_P_*, Deck perseverance (*B_P_* ∈ (−∞, +∞)); HC, Healthy controls; PC, Patient controls; SA, Suicide attempters; vSA, Violent suicide attempt; nvSA, Non-violent suicide attempt; n.s., Did not reach significance level *P* < .05. The plots display the posterior distributions of each model parameter for each group. Group differences in parameter hyperdistributions were computed using Welch’s ANOVA and Games-Howell post-hoc test. ^a^ indicates statistical significance at holm-corrected *P* < .05. ^b^ indicates statistical significance at holm-corrected *P* < .001. Across both studies, the only parameter that differed significantly (*P* < .05 after correction) in SA only was decay-rate (*K*), *i.e.*, how fast participants forget about their past choices. (**A**) SPAD (older-adults) study result. Compared with HC and PC, SA show decreased raward (*A_rew_*) and punishment (*A_pun_*) learning rates, win-frequency perseverance (*B_F_*), and increased decay-rate (*K*). (**B**) SUICIDE-DECIDE (middle-aged adults stratified by attempt method violence) study results.

### Associations with K

Women showed lower decay-rates than men (*β* = –0.24, SE = 0.06, *t_(139)_* = –4.06, *P* < .001). PC showed lower (*β* = −0.21, SE = 0.06, *t_(139)_* = −3.19, *P* = .002) and SA higher decay-rates (*β* = 0.49, SE = 0.08, *t_(139)_* = 6.45, *P* < .001) than HC. Higher Go/No-Go mean RT was associated with higher decay-rate (*β* = 0.001, SE = 0, *t_(138)_* = 4.2, *P* = .014). The SUICIDE-DECIDE sample showed higher decay-rates than SPAD (*β* = 0.26, SE = 0.06, *t_(139)_* = 4.2, *P* < .001).

### Group prediction

Including *K* as a predictor (model 2) improved fit relative to BDI only (model 1), *χ²*_(2)_ = 37.9, *P* < .001, *AIC_model1_* = 152.2; *AIC_model2_* = 118.3, *ΔAIC* = 33.9. BDI and *K* were significantly associated with group membership, but with different patterns.

Relative to HC, higher BDI was associated with an increased risk of PC (*β* = 4.35, SE = 1.10, *z* = 3.94, *P* < .001, RRR = 77.79, 95% CI [8.94, 676.60]) and SA group membership (*β* = 4.74, SE = 1.13, *z* = 4.20, *P* < .001, RRR = 114.50, 95% CI [12.56, 1044.22]). While these RRRs are large and their corresponding CIs wide, effects are directionally consistent, indicating that elevated depressive symptoms robustly distinguish clinical groups from HC.

*K* was significantly associated with SA (*β* = 1.44, SE = 0.51, z = 2.80, *P* = .005, RRR = 4.21, 95% CI [1.54, 11.49]) but not PC group membership (*β* = –0.74, SE = 0.55, *z* = –1.35, *P* = .18, RRR = 0.48, 95% CI [0.16, 1.40]). Higher *K* is associated with increased relative risk of being SA, but not PC.

Overall classification accuracy in the training sample using BDI and *K* as predictors was 76.6% (82/107 correctly classified; 95% CI [67.5%, 84.2%]; Table 3). We observed substantial agreement between predicted and observed classifications (*κ* = 0.64). Besides higher accuracy than model 1, it greatly improved PC’s sensitivity (*ΔSensitivity* = 46.5%) and SA’s specificity (*ΔSpecificity* = 19%).

**Table 3.** Prediction performance in Training and Validation sample.

|  | True<br>Positive<br>n | True<br>Negative<br>n | False<br>Positive<br>n | False<br>Negative<br>n | Sensi<br>tivity<br>% | Specifi<br>city % | Balance<br>d<br>Accurac<br>y % | PPV<br>% | NPV<br>% | PSI<br>% | AUC |
| --- | --- | --- | --- | --- | --- | --- | --- | --- | --- | --- | --- |
| **Training (n = 107)** |  |  |  |  |  |  |  |  |  |  |  |
| *Model 1 (Group ~ BDI total)* |  |  |  |  |  |  |  |  |  |  |  |
| HC | 32/35 | 62/72 | 10/72 | 3/35 | 91.43 | 86.11 | 88.77 | 76.1<br>9 | 95.3<br>8 | 71.5<br>7 | 95.5<br>4 |
| PC | 2/28 | 74/79 | 5/79 | 26/28 | 7.14 | 93.67 | 50.41 | 28.5<br>7 | 74 | 2.57 | 73.7<br>8 |
| SA | 37/44 | 42/63 | 21/63 | 7/44 | 84.09 | 66.67 | 75.38 | 63.7<br>9 | 85.7<br>1 | 49.5 | 82.7 |
| *Model 2 (Group ~ BDI total + K)* |  |  |  |  |  |  |  |  |  |  |  |
| HC | 31/35 | 63/72 | 9/72 | 4/35 | 88.57 | 87.5 | 88.04 | 77.5 | 94.0<br>3 | 71.5<br>3 | 95.8<br>3 |
| PC | 15/28 | 72/79 | 7/79 | 13/28 | 53.57 | 91.14 | 72.36 | 68.1<br>8 | 84.7<br>1 | 52.8<br>9 | 90.2<br>8 |
| SA | 36/44 | 54/63 | 9/63 | 8/44 | 81.82 | 85.71 | 83.77 | 80 | 87.1 | 67.1 | 92.1<br>7 |
| **Validation (n = 47)** |  |  |  |  |  |  |  |  |  |  |  |
| *Model 2 (Group ~ BDI total + K)* |  |  |  |  |  |  |  |  |  |  |  |
| HC | 17/17 | 30/30 | 0/30 | 0/17 | 100 | 100 | 100 | 100 | 100 | 100 | 100 |
| PC | 10/12 | 31/35 | 4/35 | 2/12 | 83.33 | 88.57 | 85.95 | 71.4<br>3 | 93.9<br>4 | 65.3<br>7 | 92.8<br>6 |
| SA | 14/18 | 27/29 | 2/29 | 4/18 | 77.78 | 93.1 | 85.44 | 87.5 | 87.1 | 74.6 | 92.5<br>3 |

Abbreviations: PPV, Positive Predictive Value (proportion of positive results that are true positives:
$\frac{n_{\text{true positives}}}{n_{\text{true positives}} + n_{\text{false positives}}}$ ); NPV, Negative Predictive Value (proportion of negative results that are true
negatives: $\frac{n_{\text{true negatives}}}{n_{\text{true negatives}} + n_{\text{false negatives}}}$ ); PSI, Prognostic Summary Index (overall certainty in positive and
negative classification: $\frac{PPV + NPV}{100}$ ); AUC, Area Under the Curve (overall discrimination ability). Model 1 and
2 were trained on the SUICIDE-DECIDE sample (n = 107). Model 2 provided the best performance, and was applied to the SPAD sample (n = 47) as an independent validation sample.

Overall classification accuracy in the validation sample was 87.2% (41/47 correctly classified; 95% CI [74.3%, 95.2%]). We observed almost perfect agreement between predicted and observed classifications (*κ* = 0.81).

## Discussion

In this cross-sectional study, we analyzed latent IGT decision-making parameters to identify mechanisms related to suicidal behavior in two groups at high suicide-risk, namely older SA and vSA. Bayesian modeling revealed higher decay-rates in both older SA and vSA, indicating that recent feedback disproportionately shaped their choices.

Decay-rate improved group classification beyond depressive symptoms alone, and enhanced discrimination between PC and SA, emphasizing its potential as a marker of suicidal vulnerability.

Decay-rate was the only parameter that differentiated older SA from PC and HC, and vSA from nvSA, PC, and HC, while it did not differ between controls suggesting a suicide-specific alteration. Consistent with previous studies,^19,21^ we did not identify suicide-specific alterations on other parameters. This specific mechanism may help explain inconsistent findings in studies relying solely on global IGT performance, as aggregate scores obscure distinct processes.^17,40^

Decay-rate was unrelated to depressive symptoms, age, cognitive flexibility, or behavioral inhibition, suggesting that it captures a distinct cognitive alteration rather than a general executive deficit. However, we observed higher decay-rates in men—who typically realize more lethal acts^41^—and prior work reported a positive association between high decay-rate and suicide attempt among participants with high childhood trauma,^21^ a significant risk factor for suicide death.^2^

This narrower temporal integration window appears consistently across prior PVL^19^ and ORL^21^ models findings, showcasing its robustness. Future work should evaluate decay-rate across multiple tasks to determine its generalizability beyond the IGT, and test it in emotionally charged contexts where suicidal crises typically unfold.^42^ The ORL model is built around a biologically grounded framework (*e.g.*, separate learning rates for gain and loss, consistent with dissociable reward and punishment systems^43^). Computational modeling paired with physiological and imaging measures such as fMRI or EEG may determine whether elevated decay-rate corresponds to known biological alterations.

Meta-analytic estimates report a mean AUC of 0.81 for suicide attempts classification models,^44^ which our model’s performance compares favorably with (all AUCs > 0.90). Most published models rely solely on internal validation, increasing overfitting risk and limiting generalizability.^44^ In contrast, our model maintained high accuracy in an independent validation sample, providing stronger evidence for its predictive utility.

Our approach also differs from most suicide-prediction models. Rather than integrating large sets of demographic, clinical, or electronic health record variables, we paired a historically associated clinical symptom with a mechanistically interpretable cognitive parameter. This parsimonious strategy enhances interpretability and translation to clinical settings while leveraging the power of multimodal data.^45^ Future work should assess decay-rate across the full suicidality continuum (*e.g.*, suicidal ideation) to clarify its predictive value.

### Theoretical and clinical implications

Here, the narrower temporal integration window reflects a tendency for recent feedback to disproportionately shape behavior, but it could also relate to the difficulty of SA to envision the consequences of killing oneself and considering alternatives^42^ (*i.e.*, narrow past and future temporal integration windows). This interpretation aligns with theories of “myopic present-focus”, in which short-term crises precipitate suicide attempts.^42^ Elevated decay-rate may be a candidate correlate of suicidal vulnerability to help stratify individuals at heightened risk. Cognitive Behavioral Therapy interventions aimed at broadening the temporal integration window (*i.e.* reducing the short-sightedness) to prevent reiterations or attempts may be more suitable.^46^

### Limitations

Our cross-sectional design prevents us from determining if decay-rate increase reflects a trait-like vulnerability or a state marker. Longitudinal studies are necessary to establish temporal precedence and predictive validity. Decay-rate may relate to other cognitive mechanisms (such as encoding efficiency) that we did not measure and could be confounding factors. Our PC group was the smallest in both samples, which may have reduced the model’s ability to distinguish them and contributed to misclassification as SA. We lacked follow-up data to determine whether these PC subsequently attempted suicide. Finally, our study focused on depressed SA and did not include other psychiatric groups at risk of suicide (*e.g.*, schizophrenia, addiction). Examining whether the decay-rate effect generalizes transdiagnostically would strengthen its robustness as a suicide phenotype.

## Conclusion

We identified a reproducible decision-making alteration—elevated decay-rate—in suicide attempters across two independent samples with the highest lethality profiles.

This short-sighted feedback integration separated attempters from control groups, and further distinguished individuals who used violent means. Decay-rate emerges as a promising candidate marker to improve risk stratification and guide future mechanistically targeted interventions.

## Data Availability

All data produced in the present study are available upon reasonable request to the authors

## Acknowledgments

The authors would like to thank Antoine GIRON, Thibault GIBERT and Bérangère GOEMAERE for data collection, Fabricio PEREIRA for implementing the task, and Rabah TAMIMOU and Leonie GAZEL for monitoring the study in Nîmes.

In Jena, we would like to thank Dr, Lydia BAHLMAN, Ms, Johanna WALTHER, Ms, Anna Karoline SEIFFERT and Ms, Anna BOLLMANN for their contributions to data acquisition. We also acknowledge support by the Open Access Publication Fund of the Thüringen Universitates- und Landesbibliothek Jena.

Finally, we would like to thank Dr Nathaniel HAINES for his insights on the models used in this study, and agreeing to share the code used in the creation of Figure 1.

